# Multivariate analysis of a missense variant in *CREBRF* reveals associations with measures of adiposity in people of Polynesian ancestries

**DOI:** 10.1101/2022.09.08.22279720

**Authors:** Jerry Z. Zhang, Lacey W. Heinsberg, Mohanraj Krishnan, Nicola L. Hawley, Tanya J. Major, Jenna C. Carlson, Jennie Harré Hindmarsh, Huti Watson, Muhammad Qasim, Lisa K. Stamp, Nicola Dalbeth, Rinki Murphy, Guangyun Sun, Hong Cheng, Take Naseri, Muagututi’a S. Reupena, Erin E. Kershaw, Ranjan Deka, Stephen T. McGarvey, Ryan L. Minster, Tony R. Merriman, Daniel E. Weeks

**Author notes:** Corresponding authors **Corresponding Authors:** Lacey W. Heinsberg, PhD, RN, Postdoctoral Scholar, Department of Human Genetics, School of Public Health, University of Pittsburgh, Public Health 3102A, 130 De Soto Street, Pittsburgh, PA 15261 USA, Daniel E. Weeks, PhD, Professor of Human Genetics and Biostatistics, Department of Human Genetics, University of Pittsburgh, Public Health 3119, 130 DeSoto Street Pittsburgh, PA 15261 USA. Equal first author contributions (Jerry Z. Zhang, Lacey W. Heinsberg). **E-mail addresses and ORCID iDs (if applicable)** Jerry Z. Zhang Lacey W. Heinsberg, Mohanraj Krishnan, Nicola L. Hawley, Tanya J. Major, Jenna C. Carlson, Jennie Harré Hindmarsh Huti Watson Muhammad Qasim Lisa K. Stamp, Nicola Dalbeth, Rinki Murphy, Guangyun Sun Hong Cheng Take Naseri, Muagututi’a S. Reupena, Erin E. Kershaw, Ranjan Deka, Stephen T. McGarvey, Ryan L. Minster, Tony Merriman, Daniel E. Weeks.

## Abstract

**Background:** The Pacific-specific minor allele of rs373863828, a missense variant in CREB3 Regulatory Factor (*CREBRF*), is associated with several cardiometabolic phenotypes in Polynesian peoples, but the variant’s function remains poorly understood. To broaden our understanding of this variant, we used joint multivariate and network analyses to examine the relationships between rs373863828 and a panel of correlated anthropometric and lipid phenotypes.

**Methods:** We tested the association of rs373863828 with a panel of phenotypes (body mass index [BMI], weight, height, HDL cholesterol, triglycerides, and total cholesterol) under a multivariate Bayesian association model in a cohort from Samoa (N = 1 632), a Māori and Pacific Island (Polynesian) cohort from Aotearoa New Zealand (N = 1 419), and the combined cohort (N = 2 976). An expanded set of phenotypes (adding estimated fat and fat-free mass, abdominal circumference, hip circumference, and abdominal-hip ratio) was also tested in the Samoa cohort (N = 1 496). Bayesian networks were learned to further understand the structure of the relationships.

**Results:** In the Samoa cohort, significant associations (log_10_ Bayes factor ≥5.0) were found between rs373863828 and the overall phenotype panel (7.97), weight (8.35) and BMI (6.39). In the Aotearoa New Zealand cohort, suggestive associations (log_10_ Bayes factor between 1.5 and 5) were found between rs373863828 and the overall phenotype panel (3.64), weight (3.30), and BMI (1.79). In the combined cohort, concordant signals with stronger magnitudes were observed. In the expanded phenotype analyses among the Samoa cohort, significant associations were also observed between rs373863828 and fat mass (5.68), abdominal circumference (5.37), and hip circumference (5.15).

Bayesian networks provided evidence for a direct association of rs373863828 with weight and indirect associations with height and BMI.

**Conclusions:** When correlation structures were considered, multivariate Bayesian analyses provided additional evidence of rs373863828’s pleiotropic effects and highlighted a strong direct effect only on weight.

## INTRODUCTION

A missense variant in the CREB3 Regulatory Factor (*CREBRF*) gene, rs373863828, has been associated with higher body mass index (BMI) in Samoan adults, with each copy of the minor allele associated with a 1.4 kg/m^2^ increase in BMI [1]. This association has been replicated in a Saipanese and Guamanian (Micronesian) cohort, a Tongan cohort, a Māori and Pacific Island (Polynesian) cohort from Aotearoa New Zealand, and a Native Hawaiian cohort [2–5]. The rs373863828 variant has also been associated with greater average body fat percentage, abdominal circumference, hip circumference, and height [1, 5–7] as well as favorable lipid profiles [1, 8] and a decreased odds of diabetes [1]. Little is understood about these complex relationships. While this allele is exceedingly rare in populations of non-Pacific ancestry (Genome Aggregation Database minor allele frequency [MAF] < 3×10^−4^), it is common in Samoan and other Polynesian populations (MAF = 0.10 to 0.26) [9–11]. Given the disproportionate burden of global obesity and related cardiovascular/inflammatory diseases (e.g., dyslipidemia, gout, chronic kidney disease) observed in Polynesian groups, an improved understanding of rs373863828 is still needed.

Of note, anthropometric and lipid phenotypes are highly correlated, making them problematic for statistical analyses, particularly in genetic association studies. Specifically, the web of correlation structures both between phenotypes and between genotypes and phenotypes can be difficult to untangle through marginal analyses (i.e., statistical modeling of a single phenotype/outcome) alone. Even more problematic for rs373863828, specifically, are the paradoxical associations observed. As highlighted above, the minor allele is simultaneously a risk factor for obesity but protective against diabetes and unfavorable lipid profiles [1]. Traditional marginal analyses considering a single phenotype simply cannot support a full understanding of the phenotypic complexity of rs373863828.

In contrast, multivariate Bayesian approaches can provide more information about the statistical dependencies within a *complete disease system* as it considers many phenotypes simultaneously through a more realistic complex “web” of relationships [12]. By considering the correlation structure between variables, multivariate approaches focus on data-driven structure discovery and can provide novel insights into both previously established and unknown relationships, including the likelihood of direct and indirect effects amongst variables of interest [12]. This information provides great value for understanding biology and disease processes, which can lead to the design of better disease control and prevention programs in future public health translation.

As such, the purpose of this study was to characterize the associations of rs373863828 with a panel of correlated anthropometric phenotypes and a set of lipid profile measurements through a powerful Bayesian multivariate framework in cohorts of individuals of Polynesian descent from Samoa and Aotearoa New Zealand.

## METHODS

### Design

This was a secondary analysis of cross-sectional, observational data collected from individuals of Polynesian descent from Samoa and Aotearoa New Zealand.

### Cohorts

The Samoa cohort consisted of 3 102 individuals of Samoan ancestry recruited in 2010 from 33 villages located across the two main islands of the Independent State of Samoa – ‘Upolu and Savai’i. Details of sample selection, genome-wide genotype and phenotype data collection, and data quality checking methods have been previously reported [1, 13]. The overall objective of the original study was to understand the genetic architecture and behavioral/environmental moderators of adiposity and related phenotypes among adults in Samoa [13] which led to the discovery of rs373863828 [1].

For this secondary data analysis, a maximally unrelated set of 1 829 individuals was selected from the Samoan cohort using PRIMUS software [14] based on a second-cousin kinship threshold to remove any potential confounding effects of kinship on the analysis. An additional 169 individuals were removed due to missingness within their phenotype panels (as the statistical approaches taken in this study require complete data for all phenotypes of interest) and 28 individuals were removed following data screening for normality and outliers (described below). The final sample size for primary multivariate analyses was 1 632 individuals (Table S1). Following identical filtering steps, the final sample size for the expanded phenotype analysis (described below) was 1 496 participants (Table S1).

The Aotearoa New Zealand cohort consisted of 2 335 individuals of Polynesian ancestry relating to nine Island Nation groups: New Zealand Māori, Cook Island Māori, Samoan, Tongan, Pukapukan, Niuean, Tahitian, Tokelauan, and Tuvaluan. This cohort was recruited across the North and South Islands of Aotearoa New Zealand, including 270 (predominantly Māori) participants recruited from the Tairāwhiti region (East Coast,

North Island) in collaboration with the Ngāti Porou Hauora Charitable Trust. Details of sample selection, genome-wide genotype and phenotype data collection, and data quality checking have been previously described [3]. Participants were recruited to better understand the genetics of gout, diabetes, and kidney disease [3, 15].

Following removal of 395 individuals due to missingness within their phenotype panels, 490 individuals due to relatedness (using the same second-cousin kinship threshold approach as above), and 31 individuals due to data screening for normality and outliers (described below), a subset of 1 419 individuals remained for multivariate analysis in the Aotearoa New Zealand cohort (Table S1).

### Phenotypes of Interest

A common set of six anthropometric and lipid profile phenotypes composed of BMI (kg/m^2^), height (m), weight (kg), HDL cholesterol (HDL-C, mg/dL), triglycerides (TG, mg/dL), and total cholesterol (mg/dL) were used within both the Samoa cohort and the Aotearoa New Zealand cohort. Cohorts were first analyzed separately, followed by a mega analysis of the combined Samoa and Aotearoa New Zealand data using the common subset of traits.

LDL cholesterol data estimated with the Friedewald formula [16] were also available. However, this marker was ultimately excluded from the analysis because (1) Friedewald-derived LDL cholesterol is linearly dependent on measured HDL, TG, and total cholesterol values and (2) the Friedewald formula is invalid for high levels of TG [17]. Therefore, inferring LDL cholesterol in this manner would have resulted in a substantial sample size reduction.

While hypolipidemic medication use was not collected for the Samoa cohort, individuals who self-reported use of heart disease medication (n=17) were excluded based on prior sensitivity analyses which revealed significant associations with cholesterol levels [1]. In contrast, for the Aotearoa New Zealand cohort, heart disease medication information was not collected, but statin and diuretic medication usage was collected. To assess the impact of medication usage on the multivariate approach, we performed a sensitivity analysis using the subset of individuals from the Aotearoa New Zealand cohort who were not on statin and diuretic medication.

In addition to the phenotypes above, additional anthropometric phenotypes – fat mass (FM, kg), fat-free mass (FFM, kg), abdominal circumference (Abd C, cm), hip circumference (Hip C, cm), and abdominal-hip ratio (AHR), were available only in the Samoa cohort and were used in an expanded Samoa cohort analysis. Fat mass and fat-free mass were estimated from bioelectrical impedance measures using equations derived from direct body composition studies of Polynesian peoples living in Aotearoa New Zealand using duel-energy X-ray absorptiometry (DXA) scans as described elsewhere [18, 19].

### Statistical Analysis

We used R statistical software as the framework for data preparation, analysis routine calling, and result reporting [20]. Within both cohorts, we adjusted phenotypes for age and sex (validated by genotyping) using ordinary linear regression models. In the Aotearoa New Zealand cohort, we also adjusted phenotypes using each individual’s first four principal components derived from genome-wide genotype data to correct for potential confounding effects of population structure between different Polynesian population sub-groups [3]. Principal components adjustment was not necessary in the Samoa cohort as all participants self-reported having four Samoan grandparents [13]; homogeneous Samoan ancestry was analytically confirmed via principal components analysis of genome-wide single nucleotide polymorphism data [1].

#### mvBIMBAM

The association of rs373863828 with the panel of phenotypes was performed using the Bayesian multivariate *mvBIMBAM* framework [21, 22]. In this framework, a global null model representing no association between phenotypes and genotype is compared with an exhaustive combination of alternative models, in which all different combinations of phenotypes are associated with the genotype. For the alternative models, the methodology splits phenotypes into all possible partitions of *U, D*, and *I*, each representing ‘unassociated’, ‘directly’, and ‘indirectly’ associated. Both directly and indirectly associated phenotypes are associated with genotype, but indirectly associated phenotypes are conditionally independent of the genotype given the presence of a directly associated phenotype in the model. The evidence against the null hypothesis is the sum of Bayes factors (BF) (log_10_ scale) of all partitions weighted by a diffuse prior [21–23]. Strong evidence of association is defined as log_10_ BF > 5; suggestive evidence is defined as 1.5 < log_10_ BF < 5; and negligible evidence is defined as log_10_ BF < 1.5. Marginal posterior probabilities of association (MPPA) are calculated by summing the marginal posterior probabilities of direct and indirect association.

The sensitivity of the Bayesian multivariate *mvBIMBAM* framework to outlier values and non-normality necessitated the normalization of phenotypes [21, 22]. Residualized phenotypes were order quantile-normalized using the *OrderNorm* function from the R package *bestNormalize* [24]. We removed observations in violation of multivariate normality at an *α* = 0.01 level based on Mahalanobis distance-based test statistics following a 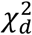 null distribution corresponding to a d-dimensional multivariate phenotype panel.

For the mega analysis, we combined the two cohorts and quantile normalized them jointly. As justified above, the Samoa cohort was adjusted for age and sex, and the Aotearoa New Zealand cohort was adjusted for age, sex, and the first four principal components estimated based on genome-wide genotype data. The residualized values were then quantile normalized jointly once again. This normalization procedure allowed for the preservation of relative ranks of observations across the two cohorts while ensuring multivariate normality.

#### Bayesian Network Analyses

We further explored the relationships between rs373863828 and the phenotypes in the quantile normalized datasets with Bayesian networks learned with the R package *bnlearn* [25, 26]. A constrained learning algorithm based on conditional independence testing (semi-interleaved HITON-PC) based on Scutari et al [27] was used to infer association and causal structure within the network [27–29]. Phenotypes and the rs373863828 were modeled as nodes with edges representing associations between nodes. We restricted the analyses so that rs373863828 could have only outgoing edges connecting to phenotypes.

The strength and directionalities of the edges of the Bayesian networks were inferred through a bootstrapped process resulting in networks that varied slightly between runs. As such, representative networks were plotted, but the quantitative strength (E_s_) and direction (E_d_) of each edge that summarized results across the total number of bootstrapped realizations was labeled on each plot. Edge strength is a measure of confidence of that edge while fixing the rest of the network structure and is defined as the empirical frequency a specific edge is observed over a set of networks learned from bootstrapped samples (i.e., the number of times the edge was present out of the total number of bootstrapped realizations). An edge was included in the network graph if its strength was larger than a significance threshold learned from the bootstrapped samples. Edge direction represents the probability of the edge’s direction conditional on the edge’s presence within the network (i.e., the number of times the edge traveled in a specific direction out of the total number of bootstrapped realizations in which it was present). Causality of arc direction should be interpreted with caution given the cross-sectional observational nature of the data [12]. However, Bayesian network analysis was designed to support causal interpretation via a set of potentially unverifiable assumptions [26, 30].

For complete analytical details, refer to documented example analysis code in the GitHub repository https://github.com/lwheinsberg/mvCREBRF.

## RESULTS

### mvBIMBAM

Using the Bayesian multivariate *mvBIMBAM* framework, we found that in the Samoa cohort the association with rs373863828, while taking the multivariate correlation structures between phenotypes (Figure 1, Figure S1) into account, was strong for the overall phenotype panel, along with the individual phenotypes weight and BMI (log_10_ BF overall = 7.97, weight = 8.35, BMI = 6.39, Table 1). There was suggestive evidence for an association with height (2.15) and negligible evidence for an association with HDL-C, TG, and total cholesterol (Table 1). The evidence of association was weaker in the Aotearoa New Zealand cohort with suggestive associations between rs373863828 and the overall phenotype panel, weight, and BMI (log_10_ BF overall = 3.64, weight = 3.30, BMI = 1.79), and negligible evidence for an association with height, HDL-C, TG, and total cholesterol (Table 1).

**Table 1.** Log_10_ Bayes Factors and marginal posterior probabilities of association (MPPA) for all cohorts – Samoa (SA, N = 1 632, Overall log_10_ BF = 7.97), Aotearoa New Zealand (A/NZ, N = 1 419, Overall log_10_ BF = 3.64), and combined (C, N = 2 976, Overall log_10_ BF = 11.34) cohorts.

|  | Weight |  |  | BMI |  |  | Height |  |  | HDL-C |  |  | TG |  |  | Chol. |  |  |
| --- | --- | --- | --- | --- | --- | --- | --- | --- | --- | --- | --- | --- | --- | --- | --- | --- | --- | --- |
|  | SA | A/NZ | C | SA | A/NZ | C | SA | A/NZ | C | SA | A/NZ | M | SA | A/NZ | C | SA | A/NZ | C |
| **Log<sub>10</sub> BF** | **8.35** | *3.30* | **11.90** | **6.39** | *1.79* | **8.16** | *2.15* | *0.81* | *3.12* | -0.01 | -0.15 | 0.10 | -0.50 | -0.30 | -0.49 | 0.83 | -0.21 | -0.43 |
| **MPPA (%)** | 100 | 99.74 | 100 | 100 | 99.17 | 100 | 99.85 | 95.22 | 99.98 | 89.11 | 76.34 | 92.01 | 58.50 | 65.36 | 33.12 | 91.77 | 69.68 | 66.80 |
| **Directly (%)** | 87.26 | 83.31 | 89.45 | 37.65 | 64.80 | 45.03 | 36.54 | 62.00 | 40.23 | 23.25 | 60.72 | 25.53 | 25.08 | 61.99 | 29.98 | 78.67 | 60.42 | 13.37 |
| **Indirectly (%)** | 12.74 | 16.42 | 10.55 | 62.35 | 34.37 | 54.97 | 63.31 | 33.22 | 59.75 | 65.86 | 15.63 | 66.49 | 33.42 | 3.36 | 3.14 | 13.10 | 9.26 | 53.43 |
| **Unaffected (%)** | 0.00 | 0.26 | 0.00 | 0.00 | 0.83 | 0.00 | 0.15 | 4.78 | 0.02 | 10.89 | 23.66 | 7.99 | 41.50 | 34.64 | 66.88 | 8.23 | 30.32 | 33.20 |

Bold log<sub>10</sub> BF's are greater than 5 (strong), italic log<sub>10</sub> BF's are between 1.5 and 5 (suggestive). BMI, body mass index; HDL-C, HDL cholesterol; TG, triglycerides; Chol., cholesterol

**Figure 1.**
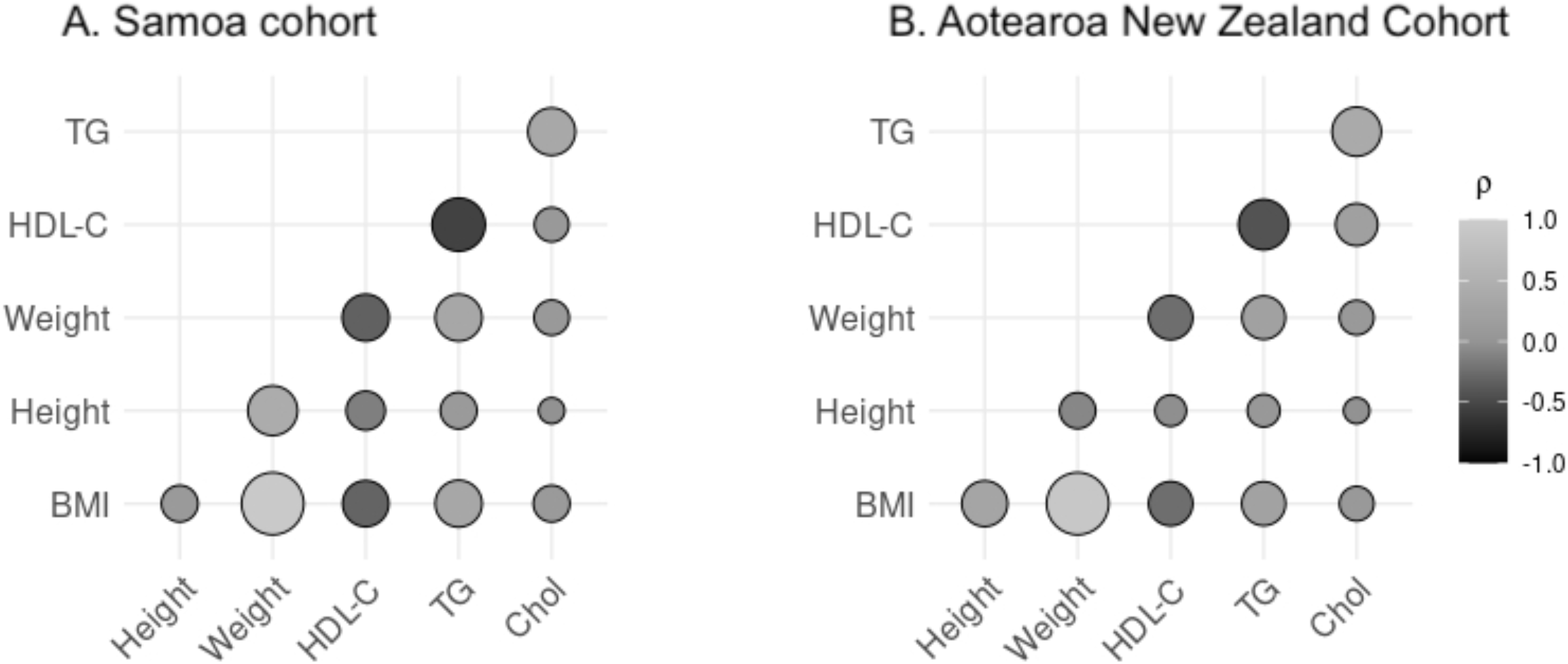
Phenotype correlation matrix for pairwise residualized phenotypes for Samoa (Left) and Aotearoa New Zealand (Right). Direction of the Pearson’s correlation coefficients (ρ) are shaded; the magnitudes are denoted by the size of the circles. See Supplemental Figure 1 for correlation matrix of expanded phenotype set. BMI, body mass index; Chol, cholesterol; TG, triglycerides; HDL-C, HDL cholesterol

When we performed a sensitivity analysis using the subset of Aotearoa New Zealand individuals who were not on statin or diuretic medication, a similar pattern of association between rs373863828 and the phenotype panel was observed, with the strongest effects noted with weight and BMI (log_10_ BF weight = 1.46, BMI = 0.90, Table S2), although all Bayes factors were below the cut-off for suggestive association (log_10_ BF < 1.5).

Within the mega analysis of the combined cohort, the evidence of association of rs373863828 with the overall phenotype panel, weight, and BMI was even greater than among the Samoa or Aotearoa New Zealand cohorts alone (log_10_ BF overall = 11.34, weight = 11.90, BMI = 8.16, Table 1). The association with height remained suggestive (log_10_ BF = 3.12), and evidence for all other associations remained negligible (Table 1).

Within the expanded anthropometric panel in the Samoa cohort, there was strong evidence for an association between rs373863828 and the overall phenotype panel, weight, BMI, fat mass, abdominal circumference, and hip circumference (log_10_ BF overall = 6.47, weight = 7.32, BMI = 5.74, fat mass = 5.68, abdominal circumference = 5.37, hip circumference = 5.15, Table 2). There was suggestive evidence for an association with height (log_10_ BF = 1.61) and fat-free mass (log_10_ BF = 1.86) and negligible evidence for associations with abdominal-hip ratio, HDL-C, TG, or total cholesterol (Table 2).

**Table 2.** Log_10_ Bayes Factors and marginal posterior probabilities of association (MPPA) for the Samoa cohort with expanded phenotype set (N = 1 496, Overall log_10_ BF = 6.47).

|  | **Weight** | **BMI** | **Height** | **FFM** | **FM** | **AHR** | **Abd C** | **Hip C** | **HDL-C** | **TG** | **Chol.** |
| --- | --- | --- | --- | --- | --- | --- | --- | --- | --- | --- | --- |
| **Log<sub>10</sub> BF** | **7.32** | **5.74** | *1.61* | *1.86* | **5.68** | 0.74 | **5.37** | **5.15** | -0.18 | -0.43 | 1.06 |
| **MPPA (%)** | 100 | 100 | 99.83 | 100 | 100 | 99.15 | 100 | 100 | 92.92 | 69.76 | 97.31 |
| **Directly (%)** | 81.01 | 31.07 | 28.50 | 40.85 | 43.97 | 24.94 | 23.98 | 27.94 | 22.20 | 36.10 | 82.88 |
| **Indirectly (%)** | 18.99 | 69.83 | 71.33 | 59.00 | 56.03 | 74.21 | 76.02 | 72.06 | 70.73 | 36.10 | 14.44 |
| **Unaffected (%)** | 0.00 | 0.00 | 0.17 | 0.15 | 0.00 | 0.85 | 0.00 | 0.00 | 7.08 | 27.80 | 2.69 |

Bold log<sub>10</sub> BF's are greater than 5 (strong), italic log<sub>10</sub> BF's are between 1.5 and 5 (suggestive). BMI, body mass index; FFM, fat-free mass; FM, fat mass; AHR, abdominal-hip ratio; Abd C, abdominal circumference; Hip C, hip circumference; HDL-C, HDL cholesterol; TG, triglycerides; Chol., cholesterol

### Bayesian Network Analyses

Bayesian networks were trained for the Samoa cohort (Figure 2), the Aotearoa New Zealand cohort (Figure 3), the combined cohort (Figure S2), and the expanded phenotype panel analysis in the Samoa cohort (Figure 4). As stated above, the strength and directionalities of the edges of the Bayesian networks were inferred through a bootstrapped process resulting in networks that varied slightly from run to run, thus representative networks are presented with quantitative edge strength:direction values that summarize results across the total number of bootstrapped realizations. In this approach, edges with high strength and strong directionality are more likely to appear in any single realization of a network, while edges with weaker directionalities may change direction across different realizations.

**Figure 2.**
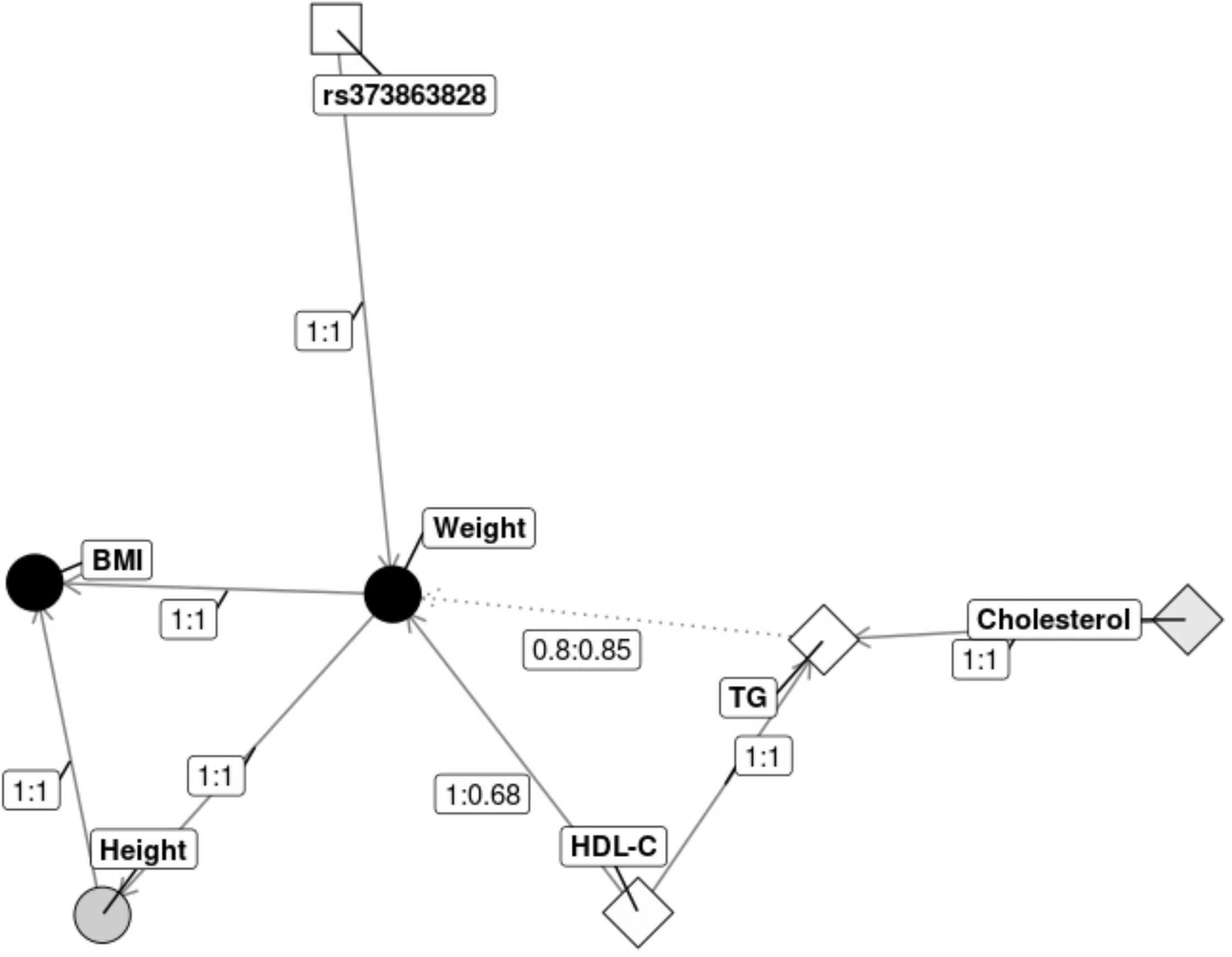
Samoa Bayesian Network. Nodes are shape coded by type (circle = anthropometric, diamond = lipids, square = the variant). Shading correspond to log_10_BF 0 (white) to 5 or greater (black). Edge labels represent (strength: directionality). Edges with strength less or equal to 0.90 are dashed. BMI, body mass index; Chol, cholesterol; TG, triglycerides; HDL-C, HDL cholesterol.

**Figure 3.**
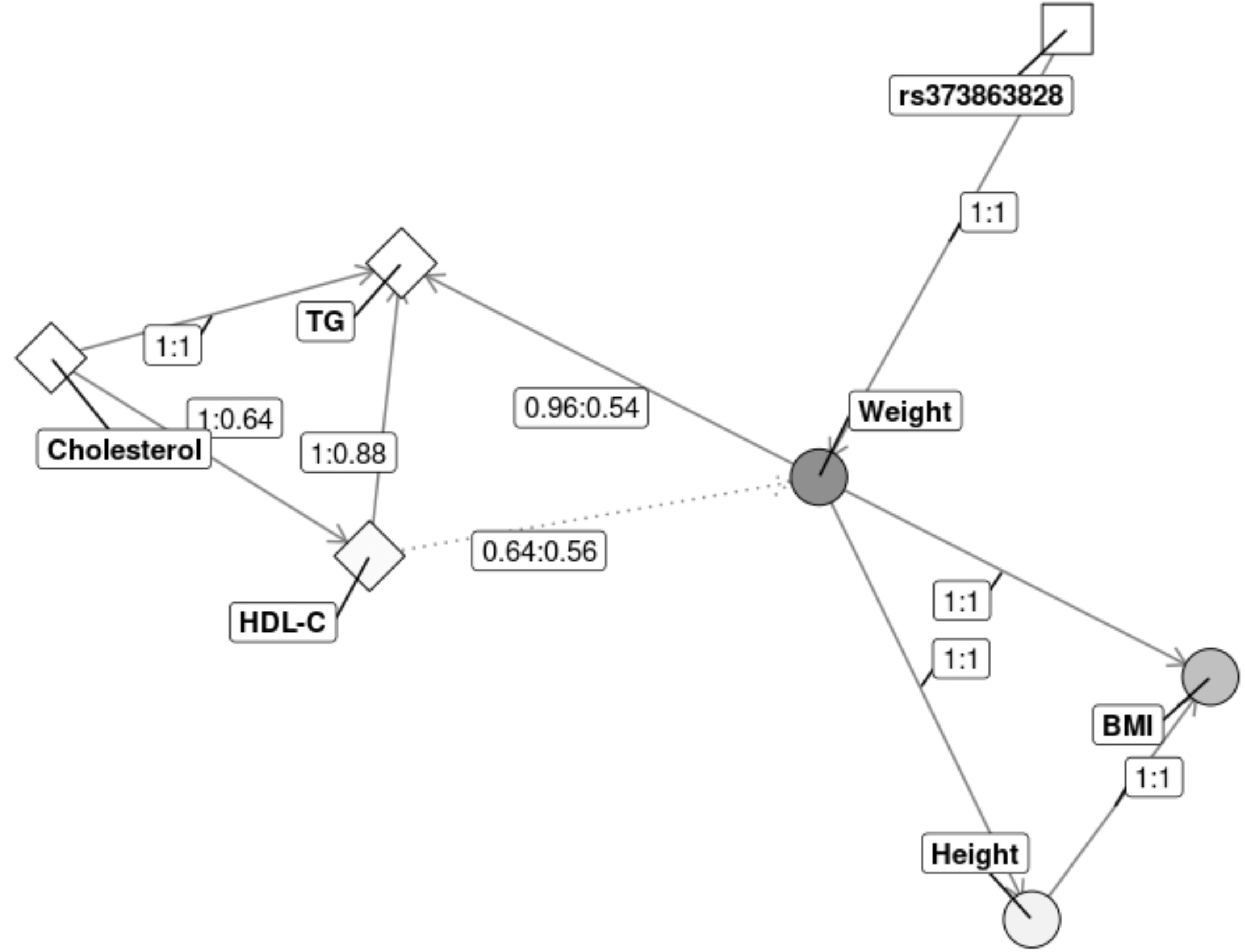
Aotearoa New Zealand Bayesian Network. Nodes are shape coded by type (circle = anthropometric, diamond = lipids, square = the variant). Shading correspond to log_10_BF 0 (white) to 5 or greater (black). Edge labels represent (strength: directionality). Edges with strength less or equal to 0.90 are dashed. BMI, body mass index; Chol, cholesterol; TG, triglycerides; HDL-C, HDL cholesterol.

**Figure 4.**
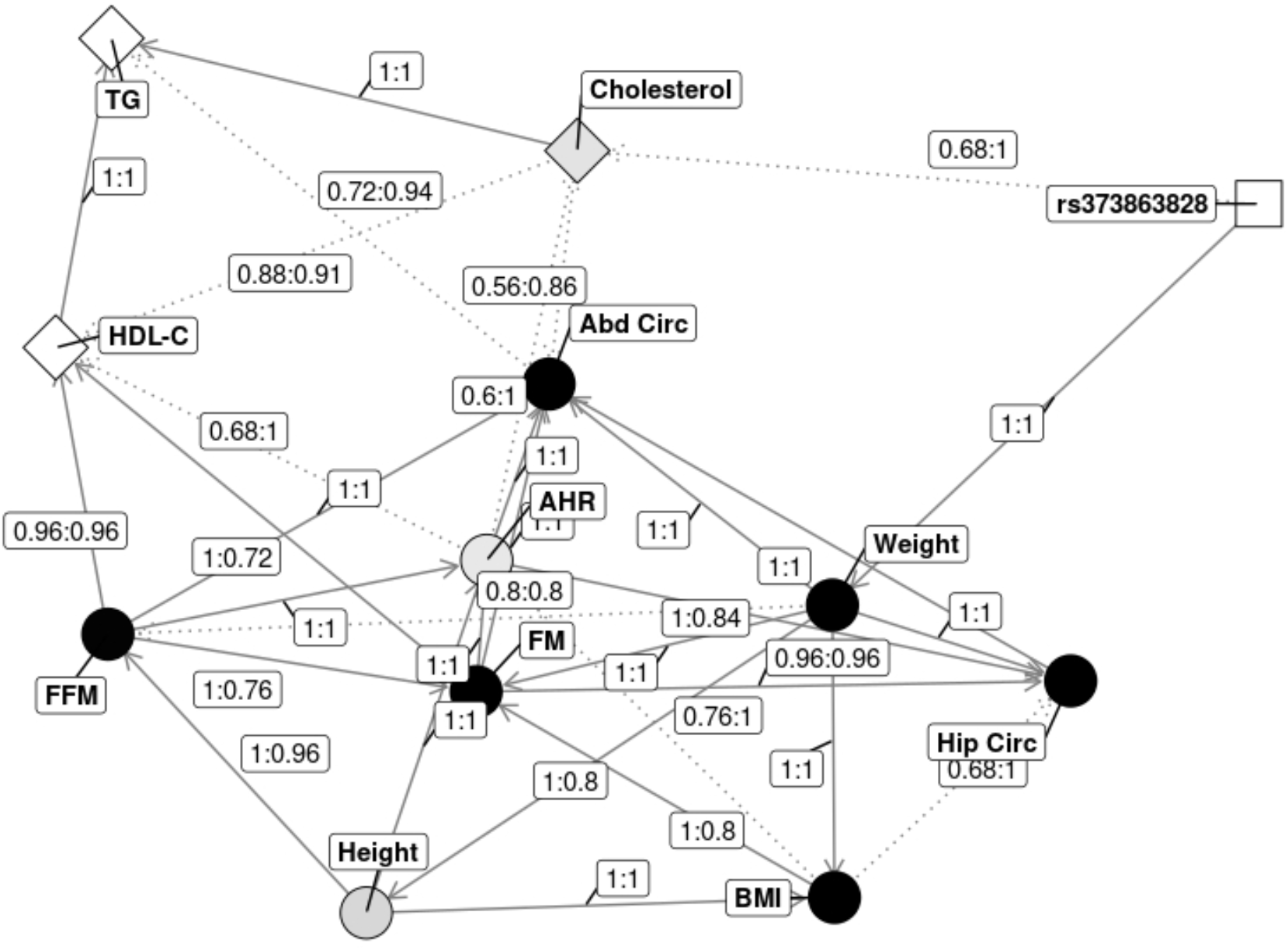
Samoa cohort Bayesian network with expanded phenotype set. Nodes are shape coded by type (circle = anthropometric, diamond = lipids, square = the variant). Shading correspond to log_10_BF 0 (white) to 5 or greater (black). Edge labels represent (strength: directionality). Edges with strength less or equal to 0.90 are dashed. BMI, body mass index; Chol, cholesterol; TG, triglycerides; HDL-C, HDL cholesterol; Abd Circ, abdominal circumference; Hip Circ, hip circumference; AHR, abdominal-hip ratio; FM, fat mass; FFM, fat-free mass.

The networks from both cohorts and the combined cohort suggested a direct association between rs373863828 and weight, indirect associations between rs373863828 and both height and BMI through weight, and a web of associations between lipid and other anthropometric traits (Figures 2-3, Figure S2). These results aligned with the evidence from *mvBIMBAM*, which suggested the association between rs373863828 and weight was more likely to be direct than indirect (probability 89% direct, 11% indirect in the combined cohort, Table 1). The inferred Bayesian networks were also consistent with *mvBIMBAM* results in the Samoa and combined cohorts, which suggested greater likelihood of indirect associations between rs373863828 and both height (probability 37% direct, 63% indirect for the Samoa cohort; 40% direct, 60% indirect in the combined cohort, Table 1) and BMI (probability 38% direct, 62% indirect for the Samoa cohort; 45% direct, 55% indirect in the combined cohort, Table 1). This was not the case in the Aotearoa New Zealand cohort, however, which resulted in more likely direct than indirect effects between rs373863828 and height (probability 62% direct, 33% indirect, Table 1) and BMI (probability 65% direct, 34% indirect, Table 1). Of note, the edge metrics for many of the relationships across all networks presented were 1:1 (i.e., consistent presence/direction in 100% of bootstrapped realizations), indicating very high statistical confidence in the strength and directions of effect (Figures 2-3, Figure S2).

In the Bayesian network analysis of the Samoa cohort with the expanded phenotype panel, an edge between rs373863828 and total cholesterol was also inferred with a moderate edge strength of 0.84 (present in 84% of the bootstrapped realizations, Figure 4). The *mvBIMBAM* results suggested a similar probability of direct association with total cholesterol (83%), though the evidence of association was negligible (log_10_ BF = 1.06, Table 2). This rs373863828-cholesterol edge was not observed in Bayesian network analyses of the two cohorts with the reduced phenotype panel (Figures 2 and 3) nor in the analysis of the combined cohort (Figure S2).

## DISCUSSION

When correlation structures were considered, multivariate Bayesian analyses provided strong evidence of the pleiotropic effects of rs373863828 including associations with weight, BMI, fat mass, abdominal circumference, and hip circumference (Tables 1, 2). Of note, rs373863828 was first discovered in a genome-wide association study of BMI [1]. Most variants that associate with increased BMI are typically associated with increased weight, but not height. Because rs373863828 is associated with increased weight [1], height [6], and BMI [1] in marginal analyses, the Bayesian approach taken here is uniquely positioned to offer probabilities of direct vs. indirect effects of rs373863828.

To that end, the strongest and most persistent direct association presented here was observed between rs373863828 and weight using both *mvBIMBAM* (log_10_ BF = 3.3 to 11.9, Tables 1-2) and *bnlearn* (Figures 2-4, Figure S2). Specifically, in the combined cohort there was strong evidence for an 89% probability of a direct effect of rs373863828 on weight. These results aligned with the relationships learned from the Bayesian networks in which 100% of bootstrapped realizations found a direct association between rs373863828 and weight across all cohorts.

The signal for association between rs373863828 and height was more varied with suggestive evidence of association in the Samoa and combined cohorts (log_10_ BF = 1.61 to 3.12, Tables 1-2) but negligible evidence of association in the Aotearoa New Zealand cohort (log_10_ BF = 0.81). In contrast to weight, in the combined cohort, the association between rs373863828 and height was favored to be indirect in both *mvBIMBAM* results (60% probability of indirect association) and *bnlearn* (100% of bootstrapped realizations suggested an indirect association between variant and height, through weight). Considered together, these results suggest that any difference in BMI due to rs373863828 genotype is most attributable to the variant’s effect on weight. This is consistent with the BMI-specific results of both *mvBIMBAM* and *bnlearn* which suggest associations between rs373863828 and BMI are most likely indirect, particularly in the Samoa and combined cohorts (Tables 1-2, Figures 2-4, Figure S2).

In the expanded phenotype panel analyses in the Samoa cohort, we observed strong evidence of association between rs373863828 and hip and abdominal circumference phenotypes (log_10_ BF abdominal circumference = 5.37, hip circumference = 5.15, Table 2), both of which are highly correlated with weight. Further, we observed stronger evidence of association between rs373863828 and fat mass compared to fat-free mass (log_10_ BF of 5.68 vs 1.86, respectively, Table 2), which contradicts prior observations that rs373863828 is associated with fat-free mass in infants [31] and adults [32], while it is associated with fat mass only in adult women [32]. While some work suggests bioimpedance-based methods do not reliably estimate fat/fat-free mass in specific ethnic groups or individuals with severe obesity states [33, 34], we have replicated work by Swinburn et al (1999) and Keighley et al (2006) that estimated fat mass of Polynesian individuals via sex, height, weight, and bioelectrical impedance measures with high accuracy [18, 19]. Specifically, in a subset of the individuals from the Samoa cohort who had both bioimpedance and DXA data available (n=424), we observed a nearly perfect correlation (r=0.97) between estimated fat mass (derived using the bioimpedance equations [18, 19]) and DXA-derived fat mass, suggesting these equations remain valid >20 years after they were derived [data not shown].

Despite the many strengths of this study, including the large sample size, complementary Bayesian analytical approaches, and focus on a historically understudied group, there are some limitations that should be acknowledged. Given that the analytical approach applied here only accepts quantitative traits, we were unable to include some important phenotypes that have been associated with rs373863828, including type 2 diabetes diagnosis [1]. While we considered analyzing a related phenotype such as fasting glucose, the addition was complicated by a substantial reduction in sample size based on the exclusion of participants by diabetes diagnosis [35] and heterogeneity of medication use and adherence [36]. Relatedly, we also examined total cholesterol, which included both detrimental LDL cholesterol and favorable HDL cholesterol. While it would be ideal to examine these values separately, the use of LDL cholesterol would have reduced our sample size substantially as described in the methods. Given the association between the rs373863828 minor allele and decreased fasting glucose [35], enhanced early insulin release [37], and favorable lipid profiles [1, 8], it will be important to examine these more analytically complex traits in the future.

In conclusion, we used two complementary Bayesian approaches to explore how rs373863828 was associated to a panel of correlated adiposity-related phenotypes when considered in a multivariate context. There was strong evidence for an association between rs373863828 and weight, BMI, fat mass, abdominal circumference, and hip circumference (Tables 1, 2), providing additional evidence of rs373863828’s pleiotropic effects. Most notably, both multivariate approaches highlighted a strong direct effect only on weight (Table 1, Figures 2-4, Figure S2), suggesting that rs373863828-specific interventions focused directly on reducing body weight might be most effective at improving other measures of adiposity and lipid profiles. In future work, it will be interesting to further examine these patterns in the context of more precise measurements of body composition and energy metabolism, as well as glucose levels, insulin release/sensitivity, and detailed lipid panels.

## Data Availability

Sample data from the Samoa cohort data are available from dbGAP, accession number: phs000914.v1.p1. Sample data from the Aotearoa New Zealand cohort are not publicly available owing to consent restrictions, but can be requested from author TRM (https://orcid.org/0000-0003-0844-8726) under an appropriate arrangement.

https://www.ncbi.nlm.nih.gov/projects/gap/cgi-bin/study.cgi?study_id=phs000914.v1.p1

## DECLARATIONS

## Acknowledgments

The authors would like to acknowledge the participants from both Samoa and Aotearoa New Zealand and all the field organizers who have made this research possible. In addition, we would like to thank the Samoan Ministry of Health, the Samoa Bureau of Statistics, and the Ministry of Women, Community, and Social Development for their continued support. We would specifically like to thank Vaimoana Lupematisila and Melania Selu for their continued engagement with the Samoan cohort. Gabrielle Sexton, Roddi Laurence, Fiona Taylor, Jordyn de Kwant, Grace Muyoma, Jill Drake, and Chris Franklin are thanked for their engagement and recruitment of Aotearoa New Zealand participants and Ria Akuhata, Nancy Aupouri and Carol Ford of the Ngāti Porou Hauora Charitable Trust for their engagement and recruitment of participants in Tairāwhiti.

This work was funded by National Institute of Health (grants R01HL093093, R01HL133040, TL1TR001858, and K99HD107030) and the Health Research Council of New Zealand (grants 08/075, 10/548, 11/1075, 14/527).

## Author’s Contributions

DEW and JZZ proposed the analysis plan. JZZ developed the analysis code, analyzed the data, and JZZ and LWH wrote the paper with guidance from DEW, as well as from JCC and RLM. MK and LWH assisted in the analyses of the data. LWH cleaned, annotated, and refined the code for public sharing on GitHub. TRM, ND, LKS, TJM, JHH, and RM contributed to collection of the Aotearoa New Zealand data. GS, HC, and RD generated the genetic marker data for the Samoa cohort. MK and TJM generated the genetic marker data for the Aotearoa New Zealand cohort. STM is the principal investigator of the Samoa cohort parent study and with DEW, was responsible for acquisition of the study funding as well as, with NLH, TN, and MSR, overseeing the 2010 Samoa data collection. All authors contributed to data interpretation, critically revised the manuscript, and approved the final version. All authors agree to be accountable for all aspects of the work.

## Code Availability Statement

To more fully document the details of our analyses, annotated analysis code, along with an example synthetic data set mirroring the statistical properties of the Samoan data set, is available in the GitHub repository https://github.com/lwheinsberg/mvCREBRF.

