## Supplementary Materials for "Multivariate analysis of a missense variant in *CREBRF* reveals associations with measures of adiposity in people of Polynesian ancestries"

### Abstract translated to Samoan

*Abstract translated to Samoan available upon request (unable to be included with preprint in accordance with medRxiv policies).*

**Figure S1.** Phenotype correlation matrix for pairwise residualized phenotypes for extended Samoa panel. Direction of the Pearson's correlation coefficients (ρ) are shaded; the magnitudes are denoted by the size of the circles.

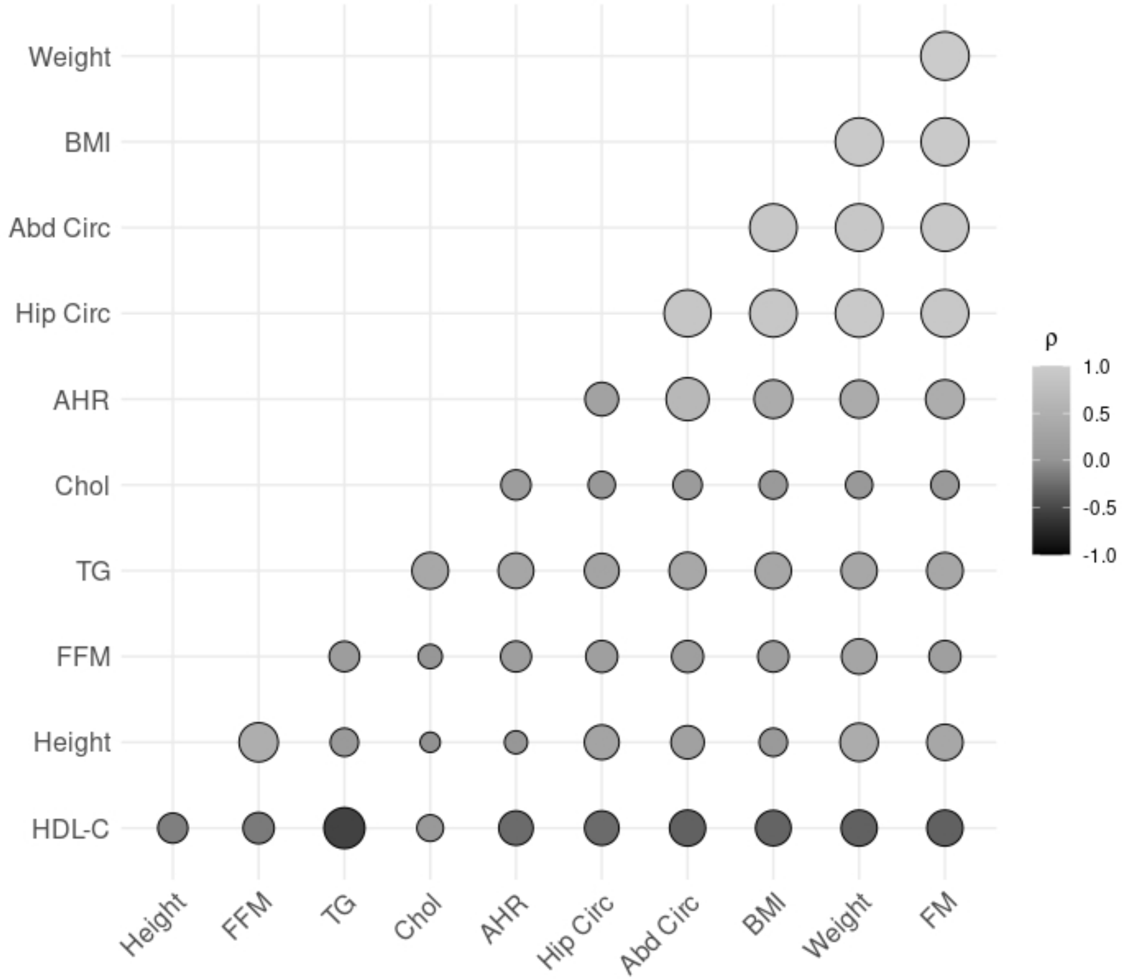


BMI, body mass index; Abd Circ, abdominal circumference; Hip Circ, hip circumference; AHR, abdomen hip ratio; Chol, cholesterol; TG, triglycerides; FFM, fat free mass; HDL-C, HDL cholesterol

**Figure S2.** Combined cohort Bayesian Network. Nodes are shape coded by type (circle = anthropometric, diamond = lipids, square = the variant). Shading corresponds to $\log_{10}$BF 0 (white) to 5 or greater (black). Edge labels represent (strength: directionality). Edges with strength less or equal to 0.90 are dashed.

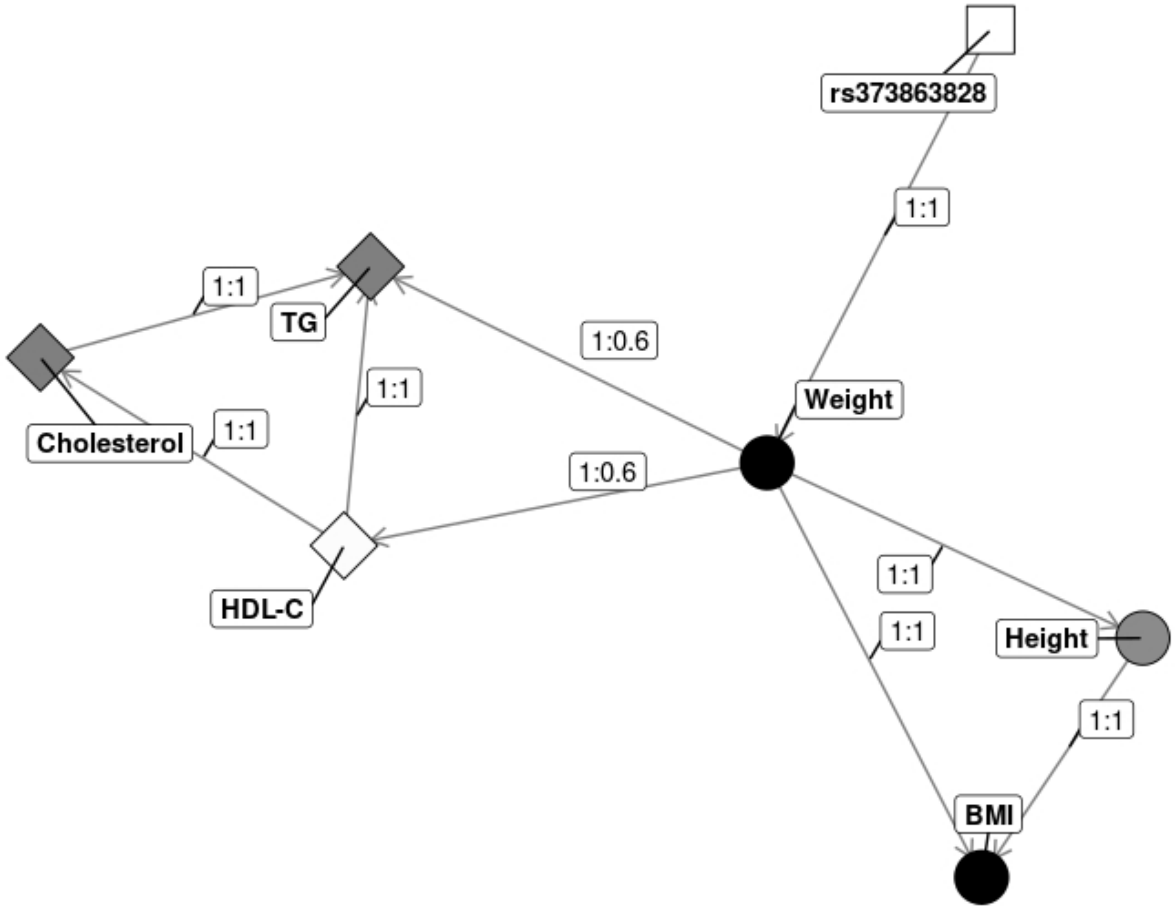

**Table S1.** Population waterfall for the Samoa and New Zealand Aotearoa cohorts.

| | **Samoa Cohort** | **Samoa Expanded Phenotypes** | **New Zealand Aotearoa cohort** | **Combined Cohort** |
|---|---|---|---|---|
| Starting sample | 3 102 | 3 102 | 2 335 | 4 890 |
| Filtering to create an unrelated set[a] | 1 829 | 1 829 | 1 450[b,c] | 3 181 |
| Filtering to remove missing phenotypes | 1 660[c] | 1 561[c] | 1 940[b] | 3 110[c] |
| Data screening (normality and outliers) | 1 632[d] | 1 496[d] | 1 419[d] | 2 976[d] |

[a]Filtering performed using the PRIMUS software based on a second-cousin kinship threshold to remove any potential confounding effects of relatedness on the analysis; [b]Due to kinship data availability restrictions, phenotype filtering was performed before relatedness filtering for the New Zealand Aotearoa cohort; [c]final sample size for each cohort prior to data screening; [d]final sample size used in multivariate analysis following data screening.

**Table S2.** Sensitivity Analysis for Aotearoa New Zealand subset (n=625) with individuals on statin and diuretics removed.

| | Weight | BMI | Height | HDL-C | TG | Chol. |
|---|---|---|---|---|---|---|
| **$Log_{10}$ BF** | 1.46 | 0.90 | 0.15 | 0.16 | -0.31 | -0.24 |
| **MPPA (%)** | 90.37 | 88.46 | 77.07 | 77.19 | 66.71 | 67.36 |
| **Directly (%)** | 62.58 | 49.41 | 38.14 | 34.80 | 26.11 | 28.36 |
| **Indirectly (%)** | 27.79 | 39.05 | 38.93 | 42.39 | 40.60 | 39.00 |
| **Unaffected (%)** | 9.63 | 11.54 | 22.92 | 22.81 | 33.29 | 32.64 |

$Log_{10}$ BF, $log_{10}$ Bayes Factor; MPPA, marginal posterior probability of association; BMI, body mass index; HDL-C, HDL cholesterol; TG, triglycerides; Chol., cholesterol
